# Effectiveness of Digital Delivery of Diabetes-Related Lifestyle Interventions in Decreasing A1c: A Systematic Review

**DOI:** 10.1101/2022.09.27.22280384

**Authors:** Hannah Rapoport

**Affiliations:** George Washington University

## Abstract

**Background:** Digital delivery of lifestyle interventions offers a potentially effective, affordable, and convenient option for patients to prevent and monitor type 2 diabetes (T2D) (Khan et al., 2019). Hemoglobin A1c (A1c) is a measure used to monitor T2D progression. Standard of care-based approaches to encourage lifestyle modification have been shown to decrease A1c, yet high healthcare costs and travel-related barriers limit the accessibility of these strategies. Rising T2D rates globally underscore the immediate need to identify the most comparably effective options that decrease A1c and address disease prevention and management.

**Objectives:** This systematic review examines the effectiveness of digital delivery of lifestyle interventions in decreasing A1c among adults globally both overall and compared to standard ofcare and monitoring only based approaches.

**Methods:** Based on application of the Navigation Guide systematic review methodology, 10 studies conducted in eight countries met the inclusion criteria and were evaluated for bias, quality, and strength of evidence. Conclusions were drawn from evaluating quantitative results.

**Results:** A systematic review of the literature demonstrated sufficient evidence of an association between digital delivery of lifestyle interventions and decreased A1c trends. Research did not show significant differences in A1c changes among the intervention groups when compared to the standard of care and monitoring only based control groups.

**Conclusions:** These results indicate that while the digital delivery of lifestyle interventions is effective in lowering A1c levels in T2D patients, these interventions do not outperform standard of care and monitoring only based approaches to prevent and manage T2D.

## Introduction

Type 2 Diabetes (T2D) rates continue to rise globally, currently affecting over six percent of the world’s population (Khan et al., 2019). Genetic predispositions, environmental exposures, and lifestyle factors contribute to T2D disease onset and long-term burden (Khan et al., 2019). Among these contributors to T2D, populations globally have the most agency over modifying lifestyle changes, including improving exercise and diet habits, to better their T2D health outcomes. Initiating and maintaining healthy lifestyle changes, however, is a challenge due to the diverse needs and socioeconomic obstacles different communities face globally (Whittemore et al., 2019). To decrease disease burden, it is crucial to address and compare culturally competent and accessible options to target lifestyle modifications with the goal of preventing and treating T2D while serving the unique needs of different populations.

Digital delivery of lifestyle interventions offers a promisingly effective, affordable, and convenient option for a wide range of patients to prevent and monitor T2D. Such interventions include, for example, mobile applications with wearable technology that target behavior change in areas such as exercise, diet, and care navigation in order to improve glycemic control.

Standard of care varies by country, but typically includes screening, diagnostic, and therapeutic actions under the direction of the patient’s physician to improve diabetes health outcomes. Diabetes Self-Management Education and Support (DSME) is a component of a typical standard of care program in different countries (Powers et al., 2017). DSME encourages self-management of diabetes by increasing patient’s knowledge of diabetes care through continued diabetes education (Powers et al., 2017). Self-monitoring of diabetes is an additional component of standard of care and includes tracking physical activity without a mobile diabetes education component and testing blood glucose daily.

The Diabetes Prevention Program (DPP) is an established lifestyle-focused alternative and/or addition to standard of care that has been systematically reviewed as a T2D care option (Knowler et al., 2002). DPP exemplifies the effectiveness of employing lifestyle interventions to delay, prevent, and improve health outcomes and lower A1c among adults with prediabetes and diabetes (Knowler et al., 2002). This program has been successfully adopted by the YMCA and is available to adults in person at YMCA centers across the United States (Rehm et al., 2017).

Hemoglobin A1c (A1c) often is used as an outcome across research to measure the effectiveness of T2D interventions, such as standard of care and DPP. A1c is a consistent metric used to measure the amount of hemoglobin with attached glucose, reflecting an individual’s average glycemic level over the past three months (Sacks, 2011). Decreased blood glucose levels and weight loss are often predictors of better T2D health outcomes, including A1c; however, measuring A1c directly is a more precise indicator of disease progression (Sacks, 2011).

Standard of care and in-person delivery of DPP to encourage lifestyle modifications have been shown to decrease A1c, yet high healthcare costs and travel-related barriers limit the accessibility of these strategies (Titus et al., 2011). Digital delivery of lifestyle interventions addresses these barriers. Thus far, researchers have not yet systematically reviewed the literature on the effectiveness of digitally based diabetes lifestyle interventions in decreasing A1c overall and compared to standard of care, including monitoring only based approaches.

A systematic review of the capability of digital delivery of diabetes-related lifestyle interventions to lower A1c is essential to an evaluation of whether this platform of delivery of lifestyle programs can effectively be offered as a treatment option for adults with T2D and prediabetes globally. It is important not only to evaluate whether digital delivery of diabetes-related lifestyle interventions effectively lowers A1c, but also whether this option is equally as effective as standard of care.

## Methods

I applied the Navigation Guide’s approach to systematically evaluate the effectiveness of digital delivery of diabetes-related lifestyle interventions on A1c level.

### Step 1. Specify the Study Question

The objective was to answer the question:

“Is digital technology as a platform to deliver lifestyle interventions to adults with T2D and prediabetes effective at lowering A1c; and if so, is this method of delivering lifestyle interventions as effective at lowering A1c compared to standard of care and monitoring-only based methods of T2D and prediabetes management?”

The PICO statement included the following:

- **Population:** Adults with T2D and prediabetes
- **Intervention:** Digital technological delivery of lifestyle interventions among adults with T2D and prediabetes globally
- **Comparator:**
  1. Care prior to intervention
  2. Standard of care and monitoring-only based methods of diabetes and prediabetes management
- **Outcome:** Lowered A1c level as an indicator of glycemic control

### Step 2: Select the Evidence

#### Search methods

I created a list of search terms to identify relevant literature by using headings and keywords. I included synonyms of key words in the search terms, as well. The search was limited by publication date; only studies published after 2010 were considered for inclusion. Search terms for each database, which include terms related to the population, intervention, comparator, and outcome are outlined in Supplemental Material, Table S1. I utilized PubMed, Web of Science, ProQuest, and Scopus to search for studies. The specific databases and numbers of papers identified per each database are outlined in Supplemental Material, Table S2. I also identified studies that were referenced in the papers identified by the search directories.

#### Study selection criteria

I selected studies in which lifestyle interventions were employed among adults with T2D and prediabetes globally. A1c decrease, as an indicator of glycemic control, was the main outcome of interest in all studies selected. Study titles and abstracts were manually screened for inclusion criteria. Studies whose titles and abstracts met the inclusion criteria were then fully screened through a full-text review. Studies were excluded if:

- The article did not contain original data or observation
- Studies were published prior to 2010
- Studies did not include randomized controlled trial elements
- Study subjects were not human
- Lifestyle interventions were employed in combination with a second intervention (ie medication)
- A1c was not the primary outcome of interest
- Study participants were under the age of 18 years
- Study participants did not have T2D or prediabetes

### Data collection

Results from each included study were aggregated into a spreadsheet for comparison. Precision of results and magnitude of effect were measured in each study using either p-values, confidence intervals, or author description, depending on the quantitative estimates presented in the studies. Study characteristics, method of administering lifestyle intervention, outcome measurement, and information used to assess risk of bias were recorded for each study and organized into a separate table.

### Step 3: Rate the Quality of and Strength of Evidence

I rated the quality and strength of the evidence in each study by assessing risk of bias (Higgins & Green 2011). I rated the quality of evidence by determining if studies should be upgraded or downgraded based on the study characteristics and determined the strength of evidence across all studies according to considerations specified in Figure 1.

**Figure 1:**
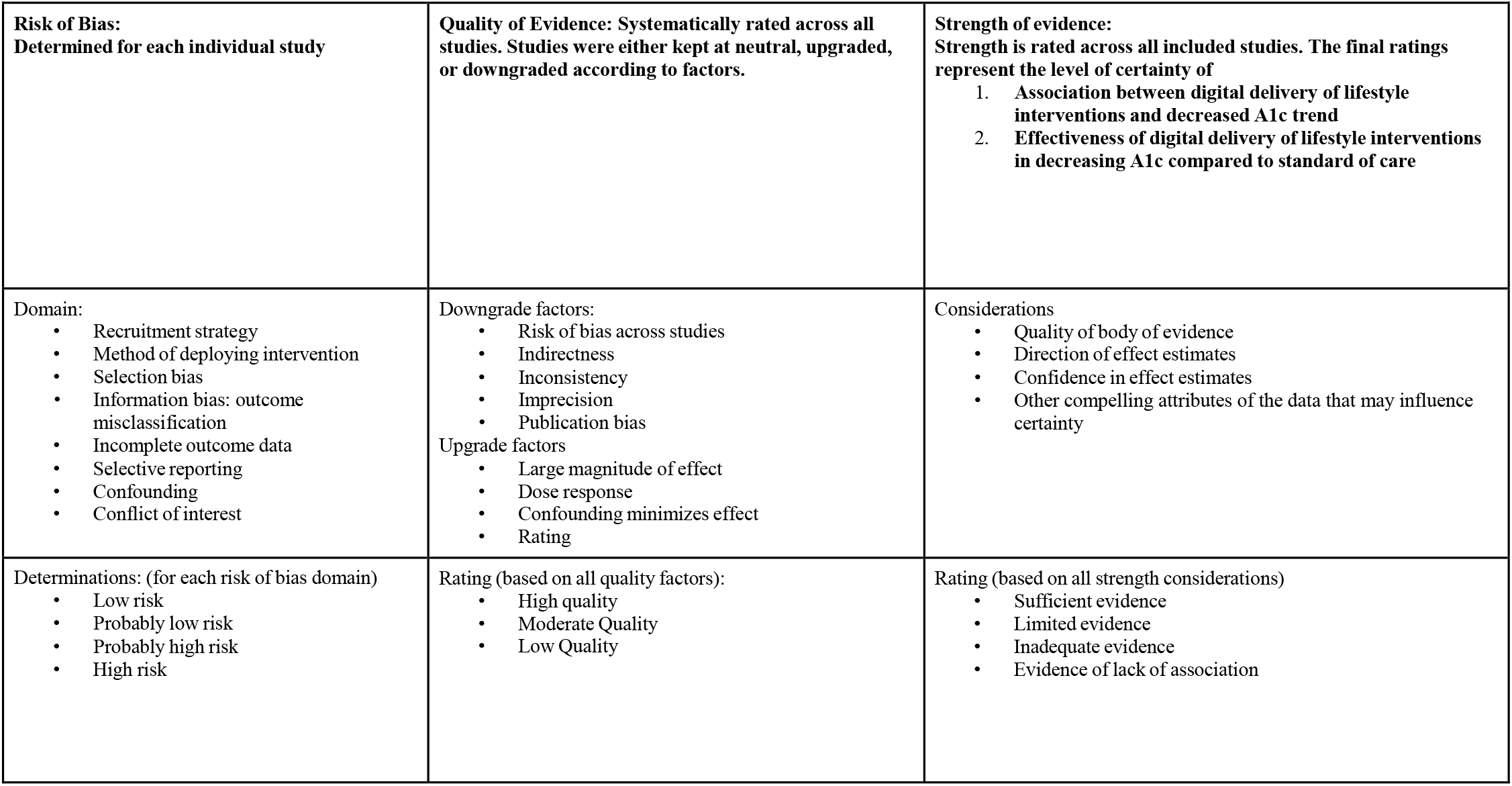
Overview of Navigation Guide Systematic review methodology used for rating the quality and strength of the human evidence.

### Assessing the risk of bias for each included study

I assessed risk of bias in the methods and data analysis sections of each study through review of each study’s recruitment strategy, inclusion of selection bias, information bias, and confounding variables. Risk of selection bias was assessed based on each study’s discussion of participant loss-to-follow-up. Use of random selection in the study design was also a key factor in determining the risk of selection bias in each study.

Information bias was evaluated by assessing A1c collection protocols from study participants. Different protocols either minimized or increased risk of non-differential and differential outcome misclassification. Evaluation of confounding variables in each study was conducted by thoroughly reading each study and determining if confounding variables were controlled for and discussed in each paper. Each study was also assessed for incomplete outcome ascertainment. Conflicts of interest among the authors of each paper were also assessed.

I assigned each risk of bias domain as “low risk,” “probably low risk,” “probably high risk,” “high risk,” or “not applicable.” The summary of each domain classification is provided in Table 1. The definition for each domain classification ensures consistency. The criterion for designating low risk of bias for confounding required that studies included randomization in the study design and statistical control of confounding variables in analysis. Studies that included randomization in the study design but did not thoroughly discuss controlling for confounding variables in analysis were found to be “probably low risk” of confounding bias.

**Table 1:** Summary of risk of bias designations for individual human studies.

|  |  |  |
| --- | --- | --- |
| **Bias** | **Rating** | **Support for judgment** |
| Recruitment Strategy | Low risk | Comprehensive recruitment strategy used uniformly across all study groups |
|  | Probably low risk | Comprehensive recruitment strategy used among most study groups |
|  | Probably high risk | Comprehensive recruitment strategy only used among one study group |
|  | High risk | Comprehensive recruitment strategy not utilized across any study groups |
| Selection bias | Low risk | Study includes random selection and accounts for Lost to Follow Up participants |
|  | Probably low risk | Study includes random selection with discussion of Lost to Follow Up but no Lost to Follow Up analysis. |
|  | Probably high risk | Study includes random selection but does not discuss Lost to Follow Up |
|  | High risk | Lost to Follow Up differs by treatment and control groups |
| Non-differential outcome misclassification | Low risk | Study took thorough measures to minimize non-differential outcome misclassification. No clear risk of non-differential misclassification of A1c. |
|  | Probably low risk | Study took some but not all measures to minimize non-differential outcome misclassification. No clear risk of non-differential misclassification of A1c; however, potential for non-differential outcome misclassification is present. |
|  | Probably high risk | Study did not take measures to prevent non-differential misclassification of A1c; however, non-differential outcome misclassification was discussed and controlled for in analysis. |
|  | High risk | Study did not take measures to prevent non-differential misclassification of A1c. Study includes clear evidence of non-differential misclassification of A1c. |
| Differential outcome misclassification | Low risk | Study took all measures to prevent differential outcome misclassification of A1c. |
|  | Probably low risk | Study took sufficient measures to prevent differential outcome misclassification of A1c. No clear risk of differential misclassification of A1c; however, potential for differential misclassification is present. |
|  | Probably high risk | Study did not take measures to prevent differential misclassification of A1c; however, differential outcome misclassification was discussed and controlled for in analysis. |
|  | High risk | Study did not take measures to prevent differential misclassification of A1c. Study includes clear evidence of differential misclassification of A1c. |
| Incomplete Outcome Data | Low risk | Complete outcome ascertainment was achieved vs. not achieved (study records are incomplete/incomplete reporting) |
|  | Probably low risk | A majority of, but not all, data was collected. Incomplete data was discussed and controlled for in analysis. |
|  | Probably high risk | Incomplete data was controlled for in analysis. |
|  | High risk | Incomplete data was not accounted for in analysis. Study made no clear efforts to achieve high data ascertainment. |

**Table 2:** Factors for evaluating the quality of the body of evidence of human studies.

|  |  |
| --- | --- |
| Evaluation Factors | Summary of Criteria |
| Downgrading factors |  |
| Risk of bias | The rating was downgraded if the study included substantial risk of bias and failed to minimize certain bias through different controls |
| Imprecise estimates | The study was downgraded if studies included few participants and few events with wide confidence intervals |
| Inconsistency | Study shows multiple conflicting results<br>No confidence intervals or p-values provided |
| Upgrading Factors |  |
| Large magnitude effect | The rating was upgraded if the exposure had a large magnitude of effect, defined as greater than 10% decrease in A1c in a from the start to the end of the study |
| Clear dose-response relationship | The rating was upgraded if the consideration of all plausible residual confounders or biases would underestimate the affect |
| Sufficiently accounted for bias | The rating was upgraded if all possible risks of misclassification of A1c were accounted for |

### Rating the quality of evidence across studies

Quality of evidence was determined by downgrade and upgrade factors. Studies were then labeled as high quality, moderate quality, or low quality. Downgrade factors included inclusion of imprecise estimates as defined by wide confidence intervals, risk of bias across studies, inconsistency, and inability to replicate study findings. Studies were upgraded if the results demonstrated a large magnitude of effect, a clear dose-response relationship, and sufficient control of confounding variables.

### Rating the strength of the evidence across studies

Strength of evidence across all studies was determined by clear indication of a trend in A1c upon exposure to digital delivery of diabetes-related lifestyle interventions, and precise estimates as indicated by narrow confidence intervals. Since this systematic review assessed whether digital delivery of lifestyle interventions affects A1c and whether this method of delivering lifestyle interventions is comparably as effective as standard of care, quantitative estimates were found in each of the studies and assessed to answer both questions. At least one quantitative estimate was found for each study (i.e. some studies had p-values but no confidence intervals).

## Results

### Included studies

The search retrieved a total of 639 unique records, of which 219 were fully screened for eligibility (Figure 2). Of these papers, 14 further met the inclusion criteria. One of these 14 articles was excluded from the study because it was based upon incomplete data collection. Three other articles were excluded because they involved single-arm study groups and did not include non-technology-based interventions as a comparator. A total of 10 articles were left for inclusion (Table 4). Quinn et al. was the only study to use a Cluster Randomized Trial study design while the remaining studies used a Randomized Control Trial study design. All studies included in this review were conducted between 2010-2018 and included populations located in eight countries. The number of participants across the studies ranged from 37-2,062 (Table 4).

**Figure 2:**
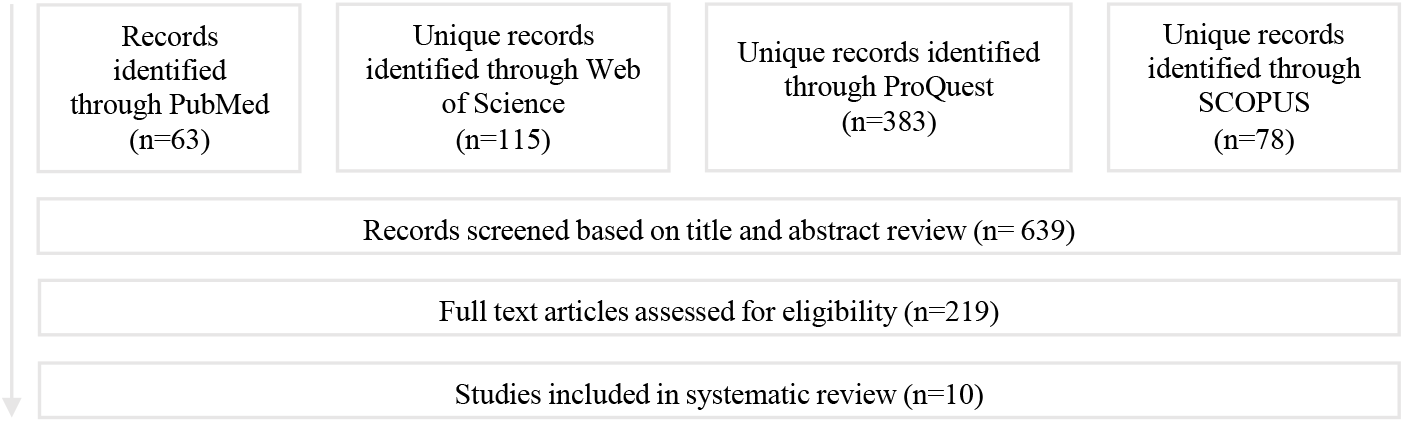
Literature Search and Screening Process.

### Risk of bias assessment for individual studies

There was generally a low risk of bias across the 10 studies (Figure 3A). According to the Navigation Guide criteria, selection bias was identified as the most common type of risk of bias within the 10 studies, while differential outcome misclassification of A1c was the least common (Figure 3B). Hilmarsdóttir et al. was among the three studies classified as having “probably high risk of selection bias.” The small-sample study had a high loss-to-follow-up (LTFU) rate, including 16.7 percent LTFU in the intervention group and 21.1 percent in the control group. Hilmarsdóttir et al. did not indicate whether the characteristics of those LTFU in the intervention group differed from those in the control group, suggesting a high potential for selection bias. Although selection bias was identified as the most common type of risk of bias across the body of evidence, there was generally still low risk of selection bias among the studies. For example, three of the 10 papers were classified as “probably low risk of selection bias,” and another four papers were classified as “low risk of selection bias.” Wayne et al., for example, discussed that although participants were lost to follow-up among both the intervention and control group, there were no statistically significant differences between the dropouts and trial completers for A1c or for demographic variables. This statistical significance gives confidence that there was low risk of selection bias in this study because the LTFU participants among both groups did not differ by outcome.

**Figure 3A:**
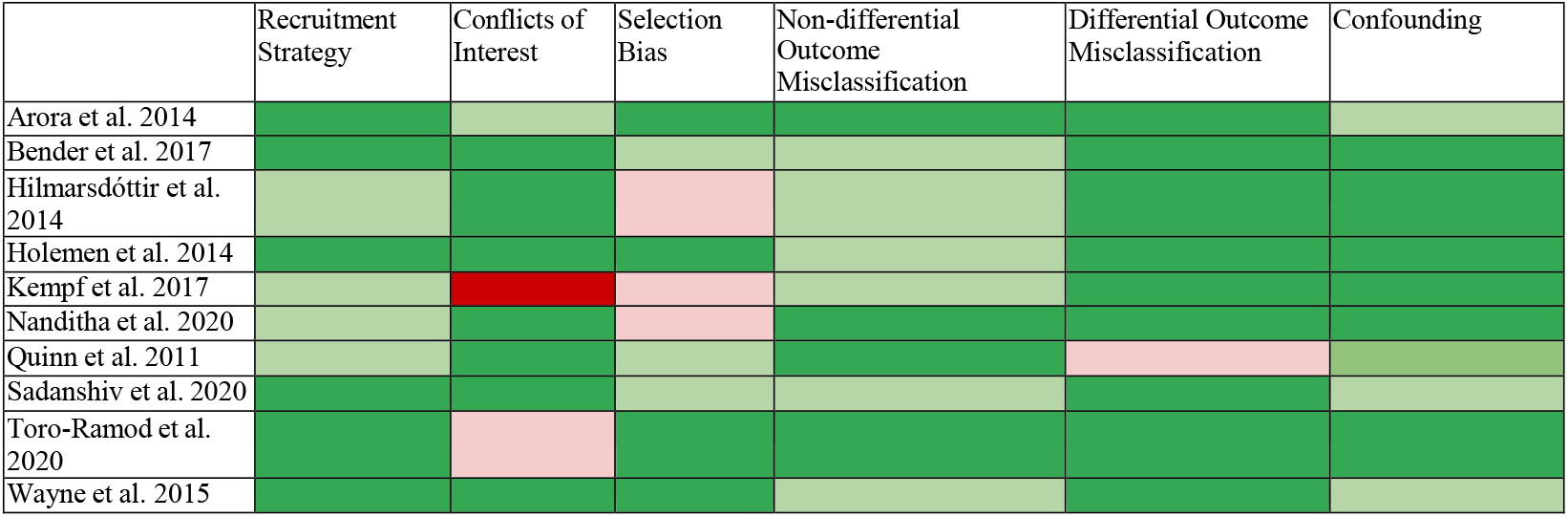
Summary of risk of bias judgments.

**Figure 3B:**
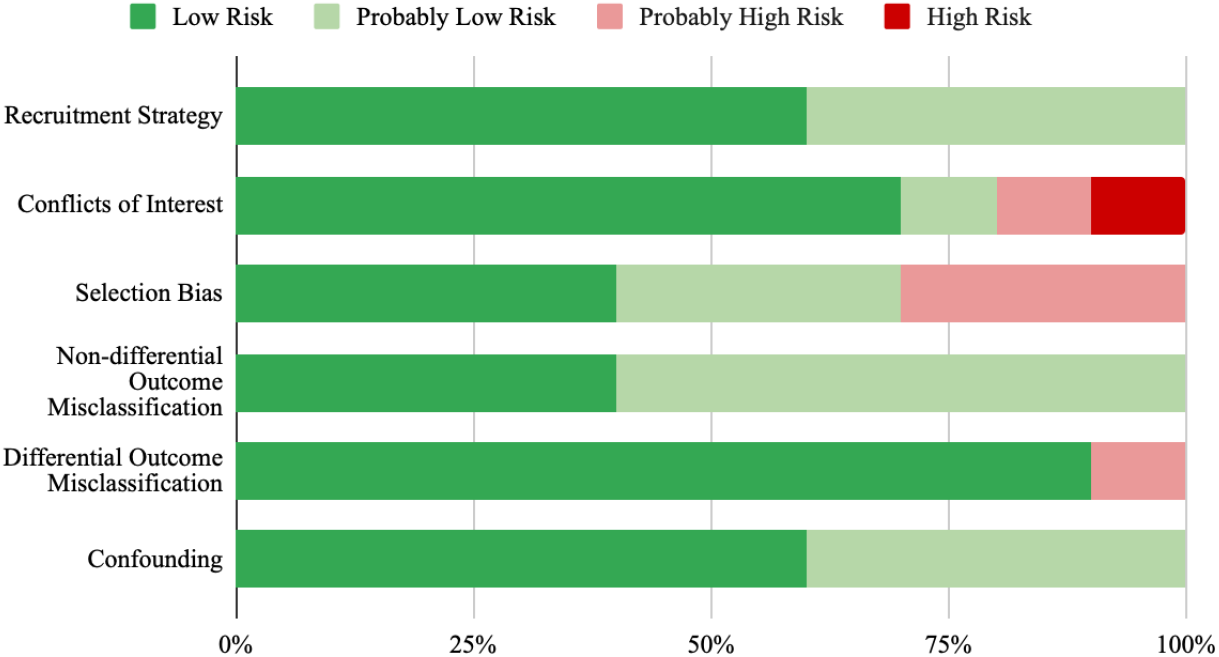
Cumulative Risk of Bias.

Given that A1c was measured uniformly among all but one of the studies, differential outcome misclassification of A1c was the least common type of risk of bias. Quinn et al. was the only study that was classified as “probably high risk” for inclusion of differential misclassification of A1c because randomization was based on site rather than by individual participants. If one site’s HbA1c calibration was off for the treatment group or intervention group, this would affect the treatment and control sites differentially. By contrast, the other nine studies uniformly measured HbA1c, so that if calibration of HbA1c measurement was off, the outcome would be non-differentially misclassified, but there would still be low risk of differential outcome misclassification of A1c.

The risk of bias in the studies’ recruitment strategies was low among a majority of the studies, considering that each study used uniform and thorough methods of recruitment across both treatment and intervention groups. A minority of studies were classified as “probably low risk” because their recruitment methods were less thorough. For example, Kempf et al. recruited participants in Germany either through attending physicians or newspaper articles. A more diverse pool of participants could have been recruited from more accessible media outlets such as Facebook or Google advertisements (Kempf et. al, 2017). Comparatively, Arora et al. used a comprehensive Emergency Department Information system to flag participants who were eligible to participate in their study.

Funding sources and declared conflicts of interest were evaluated across all studies. Of the 10 studies, Kempf et al. was the only study that included high risk of bias due to a conflict of interest, which arose from the fact that some authors were employed by the funding agency.

Across all 10 studies, there was low risk and probable low risk of confounding bias, since each of the studies took measures to prevent confounding variables in the study design and analysis. All of the studies included randomization into intervention or control groups. While Quinn et. al. randomized based on clinical site rather than randomizing participants, group assignments were concealed until a clinic agreed to be included in the study.

### Quality of the body of evidence

The study rating of human evidence on any criteria of the studies was neither downgraded nor upgraded, resulting in an overall quality of the human evidence rating of “moderate” (Table 6). A comprehensive literature search was conducted to compile studies of a wide variety of sizes and funding sources.

### Strength of the body of evidence rating

Strength of evidence considerations:

### Quality of body of evidence

Moderate quality of evidence

### Direction of effect estimate

Decreasing A1c trend among the intervention groups overall, although not all estimates were statistically significant. Result trends also indicated that A1c decreases in the intervention group were not statistically more significant than the A1c decreases in the control groups. These trends show that digital delivery of lifestyle interventions is as effective, but not more effective than standard of care approaches to lowering A1c among adults with T2D and prediabetes globally.

### Confidence in effect estimate

Further research with larger participant cohorts would likely confirm these results.

### Strength of evidence

Sufficient evidence

I compared these considerations to the definitions in Table 3 and concluded that there is sufficient evidence that digital delivery of lifestyle interventions leads to decreasing A1c trends. There is further sufficient evidence that digital delivery of lifestyle interventions is as effective at lowering A1c compared to standard of care and monitoring-only based approaches.

**Table 3:**
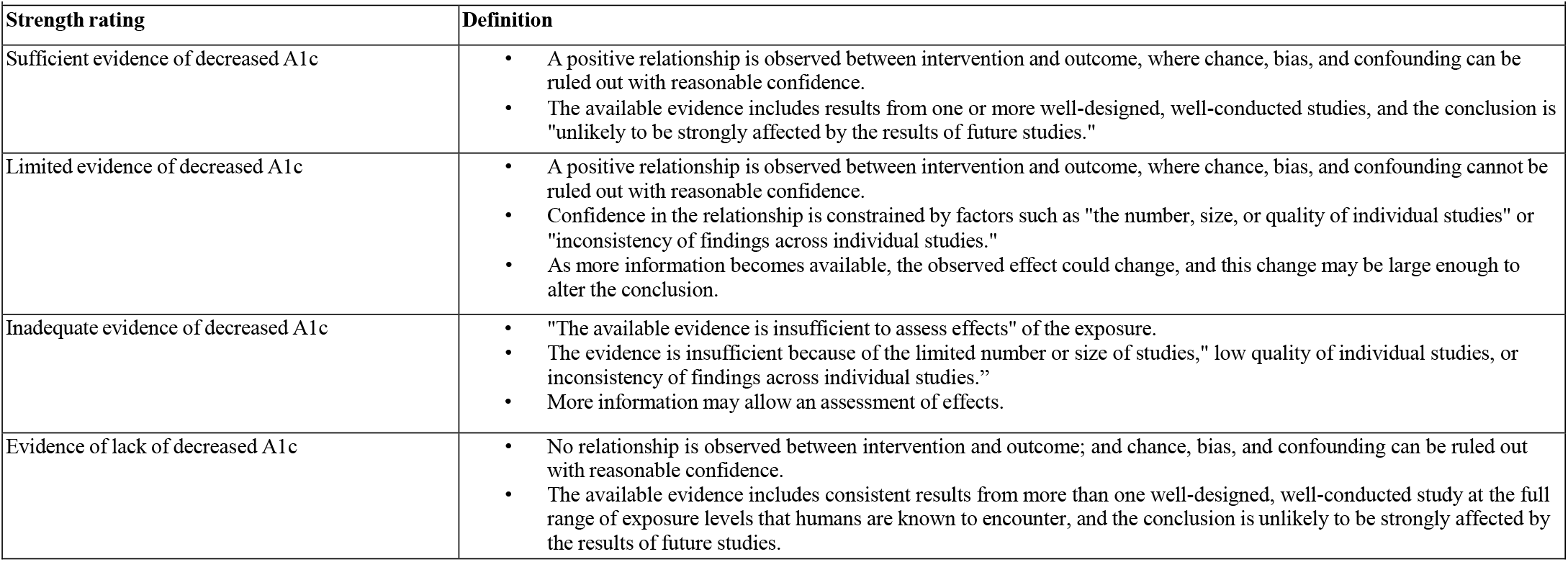
Strength of evidence definitions for human evidence (Johnson et al. 2014)

**Table 4:** Summary of Study Characteristics.

| Table 4: Summary of Study Characteristics |  |  |  |  |  |  |
| --- | --- | --- | --- | --- | --- | --- |
| Source | Number of participants | Age of Study Participants | Location/Population of Study | Study Period | Study Years | Intervention Type |
| Arora et al. 2014 | 128 | Mean at baseline 50.7 | Los Angeles, County | 6 months | Recruitment took place August 2011 and November 2011. | Text message based |
| Bender et al. 2017 | 45 participants | Mean = 58 years old (SD 10 years) | Filipino Americans in San Francisco Bay Area | 6 months | Recruitment took place between December 2014 -December 2015 | Fitbit accelerometer, mobile app with diary for health, behavior tracking, and social media (Facebook) for virtual social support groups |
| Hilmarsdóttir et al. 2020 | 37 participants | Mean = 51.2 ± 10.6 (25-70) years. | Iceland | 6 months | Data collection between March 2017 - September 2018 | Healthy-lifestyle-supporting smartphone application |
| Holman et al. 2014 | 151 participants | Mean = 57 years (SD 12) | Oslo, Norway | 12 months | Inclusion and randomization started in March 2011 and ended in September 2012. Follow-up data was finalized in October 2013. | Cell phone-based self-management system |
| Kempf et al. 2017 | 202 participants | Mean = ~60 years (Control Group: 60 [SD 8]) (Intervention Group: 59 [SD 9]) | Dusseldorf, Germany | 12 months | The first participant was enrolled on 20 February 2014; the last participant finished the 52-week follow-up on 16 December 2015. | Telemedical Lifestyle intervention Program (TeLiPro); mental motivation, a formula diet, and self-monitored blood glucose for 12 weeks. |
| Nanditha et al. 2020 | 2062 participants | Mean = 52.0 (SD 10.3) (control = 52.0; Intervention = 52.1) | India and the United Kingdom | 24 months | Recruitment in India took place between April 2012 and November 2015. Recruitment in the UK took place between June 2013 and November 2017. Follow-up was for 2 years in both countries | Cell phone SMS messages |
| Quinn et al. 2011 | 163 participants | Mean: (Group 1 = 53.2 SD 8.4) (Group 2 = 52.8 SD 8.0) (Group 3 = 53.7 SD 8.2) (Group 4 = 52 SD 8.0) Age 18–64 years (Overall 52.8) | Maryland, US | 12 months | Study was published in 2011; study does not specify which years recruitment took place | A diabetes- coaching system, using cell phones, patient/provider portals for patient-specific treatment and communication. |
| Sadanshiv et al. 2020 | 320 participants | Mean (Intervention 48.7 SD 7.4) (Control= 47.9 SD 8.0) | Healthcare workers with diabetes in, Tamil Nadu, India | 6 months | The trial recruitment started in 2015 January and ended in 2015 August. Follow-up continued until 2016 March. | Computer-generated telephonic message-based (lifestyle intervention) |
| Toro-Ramod et al. 2020 | 202 participants | Mean (Intervention = 55.7 SD 13.63) (Control group = 57.5 SD 12.45) | US adults - Stony Brook Medicine's tertiary care ambulatory clinics | 12 months | Recruitment took place between October 2016 and June 2017 (12 months of follow up) | Mobile-delivered DPP |
| Wayne et al. 2015 | 138 participants | Mean = 53.2 (SD 11.3) | Toronto, Canada | 6 months | Recruited between March 2012 – October 2013 | Mobile support and health coaching |

Results across all but one study showed a consistent downward trend of A1c among the intervention group. Although not all studies provided confidence intervals or p-values, the decrease in A1c among the intervention groups was definitely significant in six of the studies, which demonstrates the magnitude of the effect of the decrease in A1c among the participants receiving the intervention. One of the six studies, Bender et al., found that the digital intervention significantly decreased A1c at three months; however, the decrease in A1c was no longer significant at six months. This result indicates that the digital intervention may expedite A1c decrease even if the effect is not sustained longer term. Further research would be needed to confirm this finding.

The confidence intervals and p-values for seven of the ten studies indicated that the decrease in A1c among the intervention group(s) receiving digital delivery of lifestyle interventions was not significant compared to the decrease in A1c in the control group(s) receiving standard of care and monitoring only approaches. One of the seven studies, Wayne et al., demonstrated that the decrease in A1c among the intervention group was significant compared to the decrease in A1c in the control group at three but not six months.

Overall, the consistent direction of the trends across the body of evidence presented in the studies limited the risk of bias. Additionally, the thorough control of confounding variables contributes to the moderate quality of evidence provided by the body of literature.

## Discussion

Based on this first application of the Navigation Guide systematic review methodology, including comparison of quantitative results across studies (Table 5), I concluded that there was *sufficient evidence* of an association between digital delivery of lifestyle interventions and decreased A1c trends. The decrease in A1c among the intervention groups was not significant compared to the decrease in A1c among the standard of care and monitoring only based control groups, which shows that the digital delivery of lifestyle interventions is as effective but did not outperform standard of care and monitoring only based approaches to treat and prevent T2D. These conclusions were based on “moderate” quality of evidence, which included narrow confidence intervals, when available, and limited risk of bias. Further research with larger sample sizes will likely confirm these results of downward trending A1c values upon exposure to digital delivery of lifestyle interventions.

**Table 5:** Comparison of Quantitative Estimates.

| Table 5: Comparison of Quantitative Estimates |  |  |  |
| --- | --- | --- | --- |
| Study | Quantitative Estimates | Is the A1c decrease in the Intervention group significant? | Is the A1c decrease significant compared to the control group? |
| **Arora et al. 2014** | Change in A1c intervention group: -1.05<br>Change in A1c control: -0.6<br>Change in A1c compared to control group: 0.45 (p=0.230)<br>Change in A1c compared to control group among Spanish speakers: -0.80 (p= 0.025) | Yes (there was a decrease but no confidence intervals or p-values were given) | No overall<br>Yes, among Spanish speaking participants |
| **Bender et al. 2017** | Change in A1c intervention group at 3 months: -0.49 [CI: -.80 to -.21]<br>Change in A1c compared to control group at 3 months: -0.34 [CI: -0.70 to 0.04]<br>Change in A1c intervention group at 6 months: 0.15 [CI: -0.03 to 0.37]<br>Change in A1c waitlist control group receiving intervention at 6 months: -0.18 [CI: -0.42 to 0.7] | Yes at 3 months<br>Yes at 6 months (downward trend but not statistically significant) | No |
| **Hilmarsdóttir et al. 2020** | Change in A1c intervention group at 6 months: -0.7 (p<0.05)<br>Change in A1c between groups: 1.625 (P = .190) | Yes | No |
| **Holman et al. 2014** | Change in A1c among FTA intervention group at 1 year: -0.31 [CI: -0.67 to 0.05]<br>Change in A1c among FTA-HC intervention group change at 1 year: -0.15 [CI: -0.58 to 0.29]<br>Change in A1c after 1 year in control group: -0.16 [CI: -0.50 to 0.18] Decrease but not statistically significant | Yes (downward trend but not statistically significant) | No |
| **Kempf et al. 2017** | Change in A1c Intervention group at 12 weeks: -1.1 [SD +/- 1.2] (p<0.0001)<br>Change in A1c Intervention group at 26 weeks: -0.9 [SD +/- 1.2] (p<0.0001)<br>Change in A1c Intervention group at 52 weeks: -0.7 [SD +/- 1.3] (p<0.001)<br>Estimated treatment difference in the fully adjusted model: -0.8 [CI: 1.1 to 0.5] (P<0.0001) | Yes | Yes |
| **Nanditha et al. 2020** | Baseline intervention group: 6.1 [SD 0.1]<br>Change in A1c in intervention group at 6 months 0.0 [SD 0.4]<br>Change in A1c in intervention group at 12 months: -0.1 [SD 0.5]<br>Change in A1c in intervention group at 24 months: .1 [SD 0.05] | No | No |
| **Quinn et al. 2011** | Change in A1c CPDS intervention group at 12 months: -1.9 [CI -2.3 to -1.5]<br>Change in A1c Usual care intervention group at 12 months: -0.7 [CI -1.1 to -0.3]<br>Change in A1c decrease in intervention group compared to A1c decrease in control group after 12 months: 1.2 (p < 0.001) | Yes | Yes |
| **Sadanshiv et al. 2020** | Change in A1c in intervention group at 6 months: -0.54 [SD 1.09]<br>Change in A1c in control group at 6 months: -0.06 [SD 0.96]<br>Change in A1c decrease in intervention group compared to A1c decrease in control group: -0.48 [SD: 0.12] [CI: -0.71 to -0.25] (p<0.001) | Yes (downward trend but no CI was given) | Yes |
| **Toro-Ramod et al. 2020** | Change in A1c in intervention group at 6 months: -0.15 [CI: -0.22 to -0.08] (p<0.001) (Effect Size: 0.08)<br>Change in A1c in intervention group at 12 months: -0.23 [CI: -0.32 to -0.14] (p<0.001) (Effect Size: -0.13)<br>Change in A1c in intervention completers group at 6 months: -0.17 [CI: -0.25 to -0.10] (p<0.001) (Effect Size: -0.06)<br>Change in A1c in intervention completers group at 12 months: -0.28 [CI: -0.37 to -0.19] (p<0.001) (Effect Size: -0.38)<br>Difference in A1c levels between intervention and control group at 6 months: 0.004 [SE: 0.05] (p=0.94)<br>Difference in A1c levels between intervention and control group at 12 months: 0.006 [SE: 0.07] (p=0.93)<br>and 12 months:<br>Difference in HbA1c levels between Intervention and control group at 6 months: mean difference 0.004, SE 0.05; P=.94) | Yes | No |
|  | Difference in HbA1c levels between intervention and control group at 12 months: mean difference 0.006%; SE 0.07; P=.93 |  |  |
| Wayne et al. 2015 | Change in A1c in intervention group at 3 months ANOVA: -0.86 [CI: -0.47 to -1.26] (p = 0.001)<br>Change in A1c in intervention group at 6 months ANOVA: -0.84 [CI: -0.38 to -1.26] (p = 0.001)<br>Change in A1c in intervention group at 6 months: -0.82 [CI: -0.46 to -1.17] (p= 0.001)<br>Change in A1c change between intervention and control group at 3 months: 0.515 [CI: 0.500 to 0.0990] (p= 0.03)<br>Change in A1c change between intervention and control group at 6 months: 0.308 [CI: -0.180 to 0.800] (p=0.21) | Yes | Yes at 3 months<br>No at 6 months (shows that the intervention group had accelerated lowered A1c) |

**Table 6:** Summary of Qualitative Evidence.

| Table 6: Summary of Qualitative Evidence |  |  |
| --- | --- | --- |
| Quality Factor | Rating | Basis |
| **Downgrade** |  |  |
| **Risk of bias across studies** | 0 | There is no indication that there is substantial risk of bias across the body of evidence among the studies included in this systematic review |
| **Indirectness** | 0 | Each study assessed population, intervention, and outcome of interest. |
| **Inconsistency** | 0 | Results across studies were mostly consistent indicating decreased trends of A1c; although only six of the ten studies found this trend to be statistically significant. Cumulatively, the studies also showed that digital delivery of lifestyle interventions is equally as effective as standard or care and mobile-only based approaches. |
| **Publication bias** | 0 | There was no indication of publication bias considering there was a comprehensive and thorough literature search and studies were all screened for bias. |
| **Upgrade** |  |  |
| **Large magnitude of effect** | 0 | I did not consider the estimated effects large among the studies that found a statistically significant decrease in A1c among the intervention group |
| **Dose response** | 0 | There was no "incremental exposure" since all studies either included an intervention, waitlist intervention, or control group. |
| **Confounding minimizes effect** | 0 | I did not find evidence to suggest that the studies failed to account for confounding variables that would reduce effect estimates |
| **Overall quality of evidence (initial rating is "moderate")** | Moderate Quality | There were no upgrades to change the quality from the initial rating. The articles consistently indicate that mobile delivery of lifestyle interventions are associated with a downward trend of A1c; this approach is equally as, but not more effective than standard of care and monitoring only based approaches. |
| **Summary of qualitative findings** |  | Based on the analysis and interpretation of the evidence, I concluded that there is a positive, but not statistically significant association between digital, mHealth, and telehealth delivery of lifestyle interventions compared to standard of care and monitoring-only care for Type II diabetic and pre-diabetic adults. Overall, the findings across all studies suggest that digital components are as effective, but not more effective than standard of care and monitoring-only care. |
| **Strength considerations** |  |  |
| **Quality of the body of evidence** | NA | Moderate |
| **Direction of effect estimate** | NA | All but one study indicated a consistent downwards trend in A1c upon exposure to the digital and mHealth delivery of lifestyle interventions. Although only six of the studies found this trend to be statistically significant, the studies generally provided the same findings. The results further indicated that digital delivery of lifestyle interventions is equally but not more effective than standard of care and monitoring only based approaches to Type II diabetes management. |
| **Overall strength of evidence** | Sufficient evidence | Overall, the results showed that A1c trended downwards among participants with mHealth, digital, and telehealth delivery of lifestyle components although this downwards trend was not always significant. These results show that mHealth, digital, and telehealth delivery of lifestyle interventions is as effective as standard of care and monitoring only care approaches. |

Although each study took measures in the design and analysis to account for confounding variables, risk of residual confounding factors is a possibility in observational studies; however, I did not let this risk undermine the ability to make a statement about the available data. Due to randomizing intervention and control groups and controlling for confounding variables, I felt that I could rule out confounding “with reasonable confidence” (Table 3). I did not find evidence suggesting substantial residual confounding that could have explained the results.

### Significance of results

The conclusion that digital delivery of lifestyle interventions led to a downward trend and this intervention is as effective as standard of care and monitoring only based approaches is significant because although the intervention did not outperform standard of care and monitoring only based approaches, digital delivery of lifestyle interventions is an accessible, feasible, and long-term option for T2D diabetes care and prevention.

Standard of care as the main approach to treat and prevent T2D globally can be expensive for patients globally as these methods often require follow up appointments, which can lead to having to take time off of work and travel far distances (Mohan et al., 2019). Further, primary care might have long wait times for appointments and specialized physicians may not be available when needed (Titus et al., 2020). Accessibility is further limited by insurance willingness to cover visits, especially in the U.S. (Titus et al., 2020). Digital delivery of lifestyle interventions is a portable option of care and is available at any time when the patient is available. The convenience of digital delivery of lifestyle interventions expands accessibility of T2D care, management, and prevention (Iyengar et al., 2016). Further, digital delivery of lifestyle interventions is feasible in terms of cost, since access to mobile devices has expanded globally. In addition to expanding accessibility of care, digital delivery of lifestyle interventions has the potential to serve as a long-term care option for those that adhere to the treatment.

Therefore, although the intervention did not surpass standard of care and monitoring only based approaches, the accessibility, feasibility and long-term care potential of digital delivery of lifestyle interventions may lead to better adherence of diabetes management among adults with T2D and prediabetes.

### Future Research

Future research would be needed to expand understanding of the effectiveness of digital delivery of lifestyle interventions compared to standard of care and monitoring only approaches in longer term applications. Additionally, future research that includes larger sample sizes, will yield more precise estimates of the downward A1c trend among those receiving the intervention. Further, considering the decrease in A1c in the intervention group was clinically significant compared to the decrease in A1c among the control group among Spanish speaking participants in Arora et al., future research could stratify populations to determine which populations are more receptive and adherent to digital delivery of lifestyle interventions than others. Future research can also study incremental exposure levels to the intervention. For example, researchers can look into whether longer use of digital delivery of lifestyle interventions yields greater decrease in A1c, or if amount of interactivity involved in different types of digital platforms have different levels of decreased A1c.

### Strengths and limitations

A key strength of this overall systematic review is the diversity among the study participants. Considering the consistent results among these diverse populations included in the systematic review, these results are more likely to be generalizable to other populations. The thorough search process was a further strength of this systematic review. Over 600 study titles and abstracts were initially reviewed for inclusion and another 219 full text screenings were completed for study inclusion. This rigorous literature search produced a diverse batch of studies with staunch results. Another strength was the thorough evaluation of selection, confounding, and information bias through the systematic approach through the lens of the Navigation Guide. Due to the consistency of the Navigation Guide’s approach of evaluating each study’s risk of bias, each study was evaluated for bias using the same criteria.

This systematic review is limited by the available data found during the literature search, which may be insufficient in scope or may otherwise be narrow. Future reviews could be strengthened if investigators followed standardized reporting criteria guidelines, enabling improved quality assessment. For example, not all studies provided confidence intervals or p-values, which were needed to determine the magnitude of effect and significance of the decrease in A1c among the intervention group and also needed to compare the significance in decreased A1c between the intervention and control groups. Further, although the Navigation Guide provides a risk of bias tool that was used consistently and thoroughly to assess risk of bias in each study, there is no universally adopted criteria for risk of bias (Sanderson et al., 2007). An additional limitation of the body of evidence in the literature was the relatively small sample size used among the studies. All but one study had <350 participants; Nanditha et al. was the only study that included over 1,000 participants, however, this study was also the only study that did not find any decreasing A1c trend, potentially because participants were largely recruited from primary care centers. These participants could be active in the primary care system, meaning these participants already regularly participate in diabetes management and prevention. Further research with more thorough diverse recruitment methods might find different results.

## Conclusion

Digital delivery of lifestyle interventions is an innovative approach to T2D management that was systematically shown to be as effective as standard of care at lowering A1c. Although digital delivery of diabetes-related lifestyle interventions did not outperform standard of care in lowering A1c, populations may be more willing to adhere to digital diabetes-related lifestyle interventions than standard of care options due to time and cost-related barriers to care. This adherence due to improved accessibility has significant potential to improve health outcomes of individuals with T2D and offers a more equitable option to T2D management and care long- term.

## Data Availability

All data produced in the present study are available upon reasonable request to the authors

---

**Table S1:** Search Terms used in systematic literature search.

| Search | Database |
| --- | --- |
| #1 Lifestyle Intervention terms | Lifestyle intervention, digital physical activity program, apple watch, mHealth, digital lifestyle intervention, web-based Diabetes Prevention Program, eHealth Lifestyle Programs, telehealth, Fitbit. Telehealth, YMCA Diabetes Prevention Program |
| #2 Type 2 Diabetes terms | Type 2 diabetes, type 2 diabetes mellitus, T2DM, T2D, type II diabetes, adult-onset diabetes, b-cell function and lifestyle interventions, HbA1c, A1c, glycemic control |

**Table S2:** Search Results from Systematic Literature Search (main databases using search terms in Table S1 and additional websites.

| Source | Number of unique records identified | Number of unique records that fit initial inclusion criteria |
| --- | --- | --- |
| PubMed | 63 | 6 |
| Web of Science | 115 | 2 |
| ProQuest | 383 | 1 |
| Scopus | 78 | 1 |
| JSTOR | 39 | 0 |
| Total | 639 | 10 |

## Citations

Arora, S., Peters, A. L., Burner, E., Lam, C. N., & Menchine, M. (2014). Trial to examine text message–based mHealth in emergency department patients with diabetes (text-med): A randomized controlled trial. Annals of Emergency Medicine, 63(6). https://doi.org/10.1016/j.annemergmed.2013.10.012.

Bender, M. S., Cooper, B. A., Park, L. G., Padash, S., & Arai, S. (2017). A feasible and efficacious mobile-phone based lifestyle intervention for Filipino Americans with type 2 diabetes: Randomized Controlled Trial. JMIR Diabetes, 2(2). https://doi.org/10.2196/diabetes.8156

Higgins JPT, Green S, eds. 2011. Cochrane Handbook for Systematic Reviews of Interventions, Version 5.1.0 [updated March 2011]. Available: http://www.cochrane-handbook.org.

Hilmarsdóttir, E., Sigurðardóttir, Á. K., & Arnardóttir, R. H. (2020). A digital lifestyle program in outpatient treatment of type 2 diabetes: A randomized controlled study. Journal of Diabetes Science and Technology, 15(5), 1134–1141. https://doi.org/10.1177/1932296820942286.

Holmen, H., Torbjørnsen, A., Wahl, A. K., Jenum, A. K., SmÅstuen, M. C., Årsand, E., & Ribu, L. (2014). A mobile health intervention for self-management and lifestyle change for persons with type 2 diabetes, part 2: One-year results from the Norwegian randomized controlled trial Renewing Health. JMIR MHealth and UHealth, 2(4). https://doi.org/10.2196/mhealth.3882.

Iyengar, V., Wolf, A., Brown, A., & Close, K. (2016). Challenges in diabetes care: Can digital health help address them? Clinical Diabetes, 34(3), 133–141. https://doi.org/10.2337/diaclin.34.3.133.

Johnson, P. I., Sutton, P., Atchley, D. S., Koustas, E., Lam, J., Sen, S., Robinson, K. A., Axelrad, D. A., & Woodruff, T. J. (2014). The navigation guide—evidence-based medicine meets Environmental Health: Systematic Review of human evidence for PFOA effects on fetal growth. Environmental Health Perspectives, 122(10), 1028–1039. https://doi.org/10.1289/ehp.1307893.

Kempf, K., Altpeter, B., Berger, J., Reuß, O., Fuchs, M., Schneider, M., Gärtner, B., Niedermeier, K., & Martin, S. (2017). Efficacy of the Telemedical Lifestyle Intervention Program TeLiPro in advanced stages of type 2 diabetes: A randomized controlled trial. Diabetes Care, 40(7), 863–871. https://doi.org/10.2337/dc17-0303.

Khan, M. A., Hashim, M. J., King, J. K., Govender, R. D., Mustafa, H., & Al Kaabi, J. (2019). Epidemiology of type 2 diabetes – global burden of disease and forecasted trends. Journal of Epidemiology and Global Health, 10(1), 107. https://doi.org/10.2991/jegh.k.191028.001.

Knowler, W., Barrett-Connor, E., Fowler, S., Hamman, R., Lachin, J., Walker, E., Nathan, D., Watson, P. G., Mendoza, K. A., Caro, J., Goldstein, B., Lark, C., Menefee, L., Murphy, L., Pepe, C., & Spandorfer, J. M. (2002). Reduction in the incidence of type 2 diabetes with lifestyle intervention or metformin. New England Journal of Medicine, 346(6), 393–403. https://doi.org/10.1056/nejmoa012512.

Mohan, V., Cooper, M. E., Matthews, D. R., & Khunti, K. (2019). The standard of care in type 2 diabetes: Re-evaluating the treatment paradigm. Diabetes Therapy, 10(S1), 1–13. https://doi.org/10.1007/s13300-019-0573-y.

Nanditha, A., Thomson, H., Susairaj, P., Srivanichakorn, W., Oliver, N., Godsland, I. F., Majeed, A., Darzi, A., Satheesh, K., Simon, M., Raghavan, A., Vinitha, R., Snehalatha, C., Westgate, K., Brage, S., Sharp, S. J., Wareham, N. J., Johnston, D. G., & Ramachandran, A. (2020). A pragmatic and scalable strategy using mobile technology to promote sustained lifestyle changes to prevent type 2 diabetes in India and the UK: A randomised controlled trial. Diabetologia, 63(3), 486–496. https://doi.org/10.1007/s00125-019-05061-y.

Powers, M. A., Bardsley, J., Cypress, M., Duker, P., Funnell, M. M., Fischl, A. H., Maryniuk, M. D., Siminerio, L., & Vivian, E. (2017). Diabetes self-management education and support in type 2 diabetes. The Diabetes Educator, 43(1), 40–53. https://doi.org/10.1177/0145721716689694.

Quinn, C. C., Shardell, M. D., Terrin, M. L., Barr, E. A., Ballew, S. H., & Gruber-Baldini, A. L. (2011). Cluster-randomized trial of a mobile phone personalized behavioral intervention for blood glucose control. Diabetes Care, 34(9), 1934–1942. https://doi.org/10.2337/dc11-0366.

Rehm, C. D., Marquez, M. E., Spurrell-Huss, E., Hollingsworth, N., & Parsons, A. S. (2017). Lessons from launching the diabetes prevention program in a large integrated health care delivery system: A case study. Population Health Management, 20(4), 262–270. https://doi.org/10.1089/pop.2016.0109.

Sacks, D. B. (2011). A1C versus glucose testing: A Comparison. Diabetes Care, 34(2), 518–523. https://doi.org/10.2337/dc10-1546.

Sadanshiv, M., Jeyaseelan, L., Kirupakaran, H., Sonwani, V., & Sudarsanam, T. D. (2020). Feasibility of computer-generated telephonic message-based follow-up system among healthcare workers with diabetes: A randomized controlled trial. BMJ Open Diabetes Research & Care, 8(1). https://doi.org/10.1136/bmjdrc-2020-001237.

Sanderson S, Tatt ID, Higgins JPT. 2007. Tools for assessing quality and susceptibility to bias in observational studies in epidemiology: a systematic review and annotated bibliog-raphy. Int J Epidemiol 36:666–676.

Titus, S. K., & Kataoka-Yahiro, M. (2020). Barriers to access to care in Hispanics with type 2 diabetes: A systematic review. Hispanic Health Care International, 154041532095638. https://doi.org/10.1177/1540415320956389.

Toro-Ramos, T., Michaelides, A., Anton, M., Karim, Z., Kang-Oh, L., Argyrou, C., Loukaidou, E., Charitou, M. M., Sze, W., & Miller, J. D. (2020). Mobile delivery of the diabetes prevention program in people with prediabetes: Randomized controlled trial. JMIR MHealth and UHealth, 8(7). https://doi.org/10.2196/17842.

Wayne, N., Perez, D. F., Kaplan, D. M., & Ritvo, P. (2015). Health coaching reduces hba1c in type 2 diabetic patients from a lower-socioeconomic status community: A randomized controlled trial. Journal of Medical Internet Research, 17(10). https://doi.org/10.2196/jmir.4871.

Whittemore, R., Vilar-Compte, M., De La Cerda, S., Marron, D., Conover, R., Delvy, R., Lozano-Marrufo, A., & Pérez-Escamilla, R. (2019). Challenges to diabetes self-management for adults with type 2 diabetes in low-resource settings in Mexico City: A qualitative descriptive study. International Journal for Equity in Health, 18(1). https://doi.org/10.1186/s12939-019-1035-x.

